# Prediction of blood Sugar Regulation based on Healthy Boundaries, Ego Boundary and Post-trauma Growth in Patients with Diabetes

**DOI:** 10.1101/2021.05.11.21256630

**Authors:** Masoumeh Abrandabadi, Maryam Mashayekh

## Abstract

**Aims:** The aim of this study was prediction of blood sugar regulation based on ego boundary, healthy boundary and post trauma growth in patient with Diabetes.

**Methods:** For this purpose, 50 people with diabetes were selected by multistage cluster sampling. The questionnaires used in this study were the post trauma growth inventory (PGI), the ego strength (PIES), and Healthy Boundaries (HB) Questionnaire.

**Results:** Stepwise regression analysis showed that there were a significant positive relationship between blood sugar level (HbA1c) and ego strength, health boundaries and post-trauma growth (PTG).

**Conclusion:** The findings indicate a significant correlation between hyperglycemia and health boundaries, ego strength and post-traumatic growth. This means that controlling and recognizing the boundaries of mental health and post-traumatic emotions prevents high blood (HbA1c) sugar and Type 2 diabetes.

## Introduction

Diabetes is a chronic disease that physiological, cognitive, behavioral, emotional and social factors play a role in preventing, risk and regulating it and considered the epidemic of the twenty-first century. Diabetes is a huge healthcare burden worldwide. There is substantial evidence that lifestyle modifications and drug intervention can prevent diabetes, therefore, an early identification of high risk individuals is important to design targeted prevention strategies. (Hasan T. Abbas. et al, 2019)

In 2019, it is estimated that 4.2 million adults aged 20–79 years will die from diabetes, accounting for 11.3% of deaths from all causes. This is equivalent to eight deaths every minute. Almost half of these deaths (46.2%, 1.9 million) are estimated to occur in adults younger than 60 years. (Saeedi, at el 2020)

The prevalence of diabetes, chiefly Type 2 Diabetes Mellitus (T2DM), is particularly high in Iran. Few studies have been undertaken to psychosocially reviewpreventofT2DM.

Diabetes is an expensive medical problem in Iran and planning of national programmers for its control and prevention is necessary. The rates of T2DM are increasing.

It is associated with significant complications and a high cost of treatment, especially when glycemic control is poor. Despite its negative impact on health, data is still lacking on the possible biopsychosocial predictors of poor glycemic control among the diabetic population.(Sy-Cherng woon, at el 2020)

Diabetes and psychiatric disorders share a bidirectional association influencing one another in multiple ways and different patterns, like depression, anxiety, etc. (Balhara 2011)

Cognitive theories of depression have long held that the tendency to appraise stressful events in an irrational or distorted manner predisposes an individual to experience emotional and behavioral dysfunction (Beck, 1967; Haaga, Dyck, & Ernst, 1991). Cognitive theorists typically describe distorted cognitions as appraisals or conclusions mat reflect a bias in the processing of information or that are inconsistent with some commonly accepted views of reality (Alloy& Abramson, 1988; Beck, 1967). Recent evidence suggests that distorted appraisals may result in increased behavioral disability or dysfunction among physically ill individuals as well as in dysphoric mood (Christensen et al., in press; Flor& Turk, 1988; Smith, Follick, Ahern, & Adams, 1986; Smith, Peck, Milano, & Ward, 1988).

Depression and anxiety are common psychiatric complications affecting patients with diabetes mellitus. However, data on the prevalence of depression, anxiety, and associated factors among Malaysian diabetic patients is scarce. The Anxiety, Depression, and Personality Traits in Diabetes Mellitus (ADAPT-DM) study aimed to determine the prevalence of depression and anxiety, and their associated factors in the Malaysian diabetic population. (Sy-Cherng Wood, at el 2020) There is another study, Positive Psychological Interventions for Patients with Type 2 Diabetes: Rationale, Theoretical Model, and Intervention Development. (Jeff C. Huffman. et al, 2015) Based on the findings of previous study, we can say that by identifying the health locus of control and irrational health beliefs, it is possible that blood glucose level can be predicted in patients with Type II diabetes and reduced the Consequences of diabetes in people with it. (fathabadi.et al., 2018), so the main question of this research: Does blood Sugar Regulation predict on based of Healthy Boundaries, Ego Boundary and Post-trauma Growth in Patients with Diabetes?

## METHODS

The present study was descriptive in terms of collection and applied in terms of purpose.

### Statistical Society

In the present study, 50 randomly selected multistage clusters were selected from all patients with Type 2 diabetes who referred to diabetes treatment centers such as Taban Diabetes Clinic and Diabetes Association in Tehran in 1398. One year from the diagnosis of the disease. In the past, they had no other disease than diabetes, and they were over 25 years old, according to entry criteria. A list of more than 100 diabetics was compiled from the statistical population. The method of estimating the sample size is the Cochran’s formula.

### Research tools

The questionnaires used in this study were the post-trauma growth inventory (PTGI), and the ego strength psychological (PIES) and Healthy Boundaries Questionnaire (HB).

#### Ego Strength (PIES)

The PIES consists of 64 items devised by Markstrom et al. (1997) to measure the eight **ego strengths** (hope, will, purpose, competence, fidelity, love, care, and wisdom) delineated by Erikson (1964), Erikson (1985).

Strom et al. (1997) as the authors of this questionnaire examined the validity and reliability of this questionnaire. They confirmed the face validity, content and structure of this questionnaire and also reported it as 0.68 to evaluate its reliability from the Cronbach’s alpha coefficient calculation method. Altafi (2009) also reported Cronbach’s alpha of the list on an Iranian sample of 0.910 and the reliability of two scale halves of 0.77.

#### Post-trauma Growth (PTG)

Tedeschi and Calhoun (1995) developed the Post-Traumatic Growth Inventory (PTGI) to assess post-trauma growth and self-improvement a person undergoes. A 21-item scale built on the five-factor model of Tedeschi, this inventory is one of the most valid and reliable resources for evaluating personal growth that follows a stressful encounter.

Each of the 21 items falls under one of the five factors and are scored accordingly. A summation of the scores indicates the level of post-traumatic growth. The advantage of this scale is that the categorization of scores according to the five factors is suggestive of which area of self-development is predominant in us and which area might be a little behind.

#### Healthy Boundaries (HB)

20 QUESTION SELF ASSESSMENT FOR HEALTHY BOUNDARIES Copyright 1999. Dr. Jane Bolton, a marriage and family therapist, master results coach and contemporary psychoanalyst and is dedicated to supporting people in the fullest expression of their Authentic Selves. This includes Discovery, Understanding, Acceptance, Expression, and Empowerment of the Self.

### Executive method

To collect information, after selecting samples, before giving questionnaires to diabetic patients A brief explanation of the purpose of the research, the need for their sincere cooperation to advance The objectives of the research and the way of answering the questions were given and it was stated that only your real attitude is the case It is an opinion and there is no right or wrong answer. It was also emphasized that there is no need to mention your name and identity not enough time was given to patients to answer the questions the type of bias was the order of questionnaires.

### Analysis Method

In this research, considering that it is a correlation type of descriptive statistical methods (distribution table Frequency, central tendency indicators …) as well as inferential statistical methods including correlation coefficient Pearson and multiple regressions were used simultaneously.

### Ethical Consideration

1. The purpose of the study was explained to all participants.
2. Conscious consent was obtained from the participants.
3. Participants were assured that all data would remain confidential and that information would be kept confidential will be published collectively.
4. Participants were reassured about the optionality of participating in the study.
5. Participants were explained that they would be provided with research results if they wished. 6-Participants were assured that the names of the participants would not be mentioned in all documents related to the research and its publications.

This study was approved in the Research Ethics Committee at the Research Institute of Shahid Beheshti University of Tehran with the code IR.SBU.Rec.1398.022

## RESULTS

### Describing respondents based on demographic variables

Demographic information in this study includes gender, age, level of education, and peer housing. The following is the frequency and percentage of demographic variables in different sections. According to Table 1, 74% of the total participants in the study (50 people) were “women” and 26% were “men”. The age of 44% was “less than 50 years old” and 56% was “less than 50 years old”. The education rate was 30% for “diploma and sub-diploma”, 4% for “associate”, 48% for “bachelor’s degree”, 16% for “master’s degree” and 2% for “doctorate and higher”. 22% of people live “alone” and 28% live with “spouse”, 10% with “children” and 40% with “spouse and children”.

**Table 1:** Description of respondents in terms of demographic variables

|  | **Variable** | **Frequency** | **Percent** | **Cumulative Percent** |
| --- | --- | --- | --- | --- |
| **Sex** | F | 37 | 74 | 74 |
|  | M | 13 | 26 | 100 |
| **Age** | <50 | 22 | 44 | 44 |
|  | >50 | 28 | 56 | 100 |
| **Educe** | D | 15 | 30 | 30 |
|  | As | 2 | 4 | 34 |
|  | B | 24 | 48 | 82 |
|  | Ms | 8 | 16 | 98 |
|  | PhD | 1 | 2 | 100 |
| **live/w** | Alone | 11 | 22 | 22 |
|  | Sp | 14 | 28 | 50 |
|  | Ch | 5 | 10 | 60 |
|  | S &Ch | 20 | 40 | 100 |

### Descriptive statistics of research indicators

Table 2 lists the descriptive statistics of the research variables, including the number of respondents, the lowest value, the highest value, the mean, and the standard deviation.

**Table 2:** Descriptive statistics of research variables

|  | N<br>Statistic | Minimum<br>Statistic | Maximum<br>Statistic | Mean<br>Statistic | Std. Deviation<br>Statistic |
| --- | --- | --- | --- | --- | --- |
| PTG | 50 | 41.00 | 100.00 | 73.2000 | 13.47560 |
| PIS | 50 | 184.00 | 287.00 | 232.7200 | 31.39546 |
| HB | 50 | 29.00 | 79.00 | 52.8200 | 13.00344 |
| HbA1c | 50 | 6.00 | 9.00 | 7.3200 | .89625 |
| Valid N (listwise) | 50 |  |  |  |  |

**Table 3.** Evaluation of the reliability of the questionnaire

| **Variables** | **N of items** | **Cronbach's Alpha** |
| --- | --- | --- |
| PTG | 21 | 0.827 |
| PIES | 64 | 0.917 |
| HB | 20 | 0.867 |

To calculate the mean, we add the data of a variable and divide it by the number of observations. To calculate the standard deviation, we add the square of the distance of all values from the mean 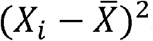 and divide the result by the number of observations minus 1 and subtract from the resulting number.

Descriptive statistics are the properties of a data set; it describes the data. Descriptive statistics are used before formal inferences are made (Evans *et al*., 2004). The data set comes from a sample. A sample comes from the population.

Inferential statistics is defined as using the sample descriptive statistics to make an inference (estimation) of the population. The sample is the observation; the estimated population is the inferred value without observation.

### Inferential analysis of the findings

The role of descriptive statistics is to collect, summarize, and describe quantitative information from samples or communities. But the researcher usually does not end his or her work by describing the information, but tries to generalize what he or she has learned from the sample group survey to larger similar groups. On the other hand, in most cases it is impossible to study all members of a community. Therefore, the researcher needs methods that can be used to generalize the results of the study of small groups to larger groups. The ways in which the characteristics of large groups are inferred based on the measurement of the same characteristics in small groups are called inferential statistics.

Statistical assumptions are claims about one or more populations that may be true or false. In other words, a statistical assumption is a claim or statement about the distribution of a population or the distribution parameter of a random variable. Statistical hypothesis is the starting point of the hypothesis test and it is basically difficult to perform a test without having statistical hypothesis. Statistical hypotheses are expressed as two types of null hypotheses (H0) and opposite assumptions (HA). The hypothesis that is tested in statistical tests is zero hypotheses, which always indicates that there is no difference. But the assumption is contrary to the same research hypothesis that can be directional or non-directional. Of course, the choice of directional hypothesis is not arbitrary and random, but research hypotheses can be developed if the previous theory or research provides evidence for it.

### Questionnair e reliability r eview

Reliability is one of the technical characteristics of measuring instruments. This concept deals with the extent to which measuring tools produce the same results under the same conditions. Definitions for reliability include those defined by Abel and Frisbee (1989): “The correlation between a setoff scores and another set of scores in an equivalent test obtained independently of a group of subjects is.”

Due to this, the range of reliability coefficient usually varies from zero (non-communication) to +1 (full communication). The reliability coefficient indicates the extent to which the measuring instrument measures the stability of the subject or his / her variable and temporary characteristics. Various methods are used to calculate the reliability coefficient of the measurement tool. Among them is Cronbach’s alpha method, which is described below.

Cronbach’s alpha is thus a function of the number of items in a test, the average covariance between pairs of items, and the variance of the total score. A value of zero for this coefficient indicates unreliability and +1 indicates complete reliability. As usual values greater than 0.7 for this coefficient can confirm the reliability of the questionnaire (Momeni and FaalQayyumi, 1396).

Since the value of Cronbach’s alpha coefficient in all questionnaire factors is greater than 0.7, Therefore, the factors of the questionnaire are at a very good level in terms of reliability. So, the reliability of the factors of the questionnaire and all the questions of the questionnaire is confirmed.

The aim of the study was to “predict blood glucose regulation (HbA1c) based on healthy boundaries, ego boundary and PTG in patients with diabetes”. According of the study the normality test for the collected data should be performed to use the appropriate test to evaluate the hypotheses.

Normal distribution means that the distribution of variables on both sides of the mean is the same, so that the distribution diagram has a bell shape. The distribution of variables is normal. Parametric tests are used to test the hypotheses, otherwise non-parametric tests are used.

According to table 4, although the significance value of Kolmogorov-Smirnov test for some variables is less than 0.05, the values of skewness and elongation for all research variables are in the range (2 and 2-). The normality of the data for these variables is confirmed. Therefore, we use parametric tests to test the research hypotheses. In total, the absolute value of the skewness and kurtosis coefficient greater than 2 indicates a violation of the normality of the data is problematic in data analysis and creates a serious problem he does. We also see in the quantitative diagrams of normal variables that all points are on a hypothetical line.

**Table 4.** Normality test results for research variables

| **Variables** | **df** | **Statistic Kolmogorov-Smirnov<sup>a</sup>** | **Sig. Kolmogorov-Smirnov<sup>a</sup>** | **Skewness** | **Kurtosis** |
| --- | --- | --- | --- | --- | --- |
| PTG | 50 | 0.079 | 0.2 | -0.393 | -0.207 |
| PIES | 50 | 0.12 | 0.07 | 0.024 | -1.329 |
| HB | 50 | 0.086 | 0.2 | 0.27 | -0.919 |
| HbA1c | 50 | 0.139 | 0.016 | 0.476 | -0.766 |

a. Lillifors Significance Correction

#### Correlation

Most of the time, researchers want to know what the relationship is between two or more variables. Correlation is the measure of the linear relationship between variables. Note that two variables may be related; but this relationship is not linear. To find the correlation between the two variables, we decide which method to use according to the type of variable being studied. We use Pearson correlation when both of our variables are quantitative (continuous) and follow a normal distribution. If even one of the variables does not follow the normal distribution, we use Spearman correlation coefficient. The value of the correlation coefficient varies between -1 and +1. A value of zero indicates that there is no linear relationship between the variables. According to the correlation matrix, if the significance value for the two indices is less than 0.05, it means that the correlation coefficient between these two indices is significant and the two indices have a high correlation (Sadegh pourGildeh and Moradi, 2013).

The partial correlation coefficient indicates the linear relationship between the two variables and the control of the effect of one or more other variables. In other words, the correlation coefficient of the two variables is in the presence of other variables. This correlation coefficient is most often used when we want to know which variable is more effective alone than other variables. In the present study, partial correlation was used to investigate the relationship between the dependent variable “HbA1c” and each of the independent variables “post-trauma growth”, “ego strength” and “healthy boundaries”. According to the correlation table, if the significance value for the two variables is less than 0.05, it means that the correlation coefficient between these two variables is significant and the two variables have a high correlation (SadeghpourGildeh and Moradi, 2013).

According to table 5, the correlation coefficient between the dependent variable “HbA1c” and each of the independent variables “post-traumatic growth”, “ego strength” and “healthy boundaries” is significant, because the significance value corresponding to these coefficients is less than 0.05 has been obtained.

**Table 5.** Correlation coefficients of research variables

| Variable |  | HbA1c |
| --- | --- | --- |
| PTG | Correlation | 0.338 |
|  | Significance (2-tailed) | 0.019 |
| PIES | Correlation | 0.332 |
|  | Significance (2-tailed) | 0.021 |
| HB | Correlation | 0.304 |
|  | Significance (2-tailed) | 0.036 |

To test the research hypotheses, regression was used, which is the regression model of the research as follows:

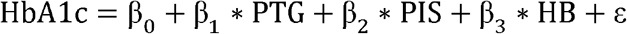

To examine the relationships between the research variables, the graph of the relationship between the dependent variable “HbA1c” and the independent variables of the research is as follows:

From the residual distribution graph and the predicted values (Figure 5), it can be seen that there is no definite relationship between the residuals and the predicted values, which is consistent with the assumption of linearity. Also, from the normal quadratic diagram for the residues, it can be seen that the residues are relatively normally distributed. Because according to this diagram, if all the points on the bisector are in the first quarter, then the remainder completely follows the normal distribution.

**Figure 1.**
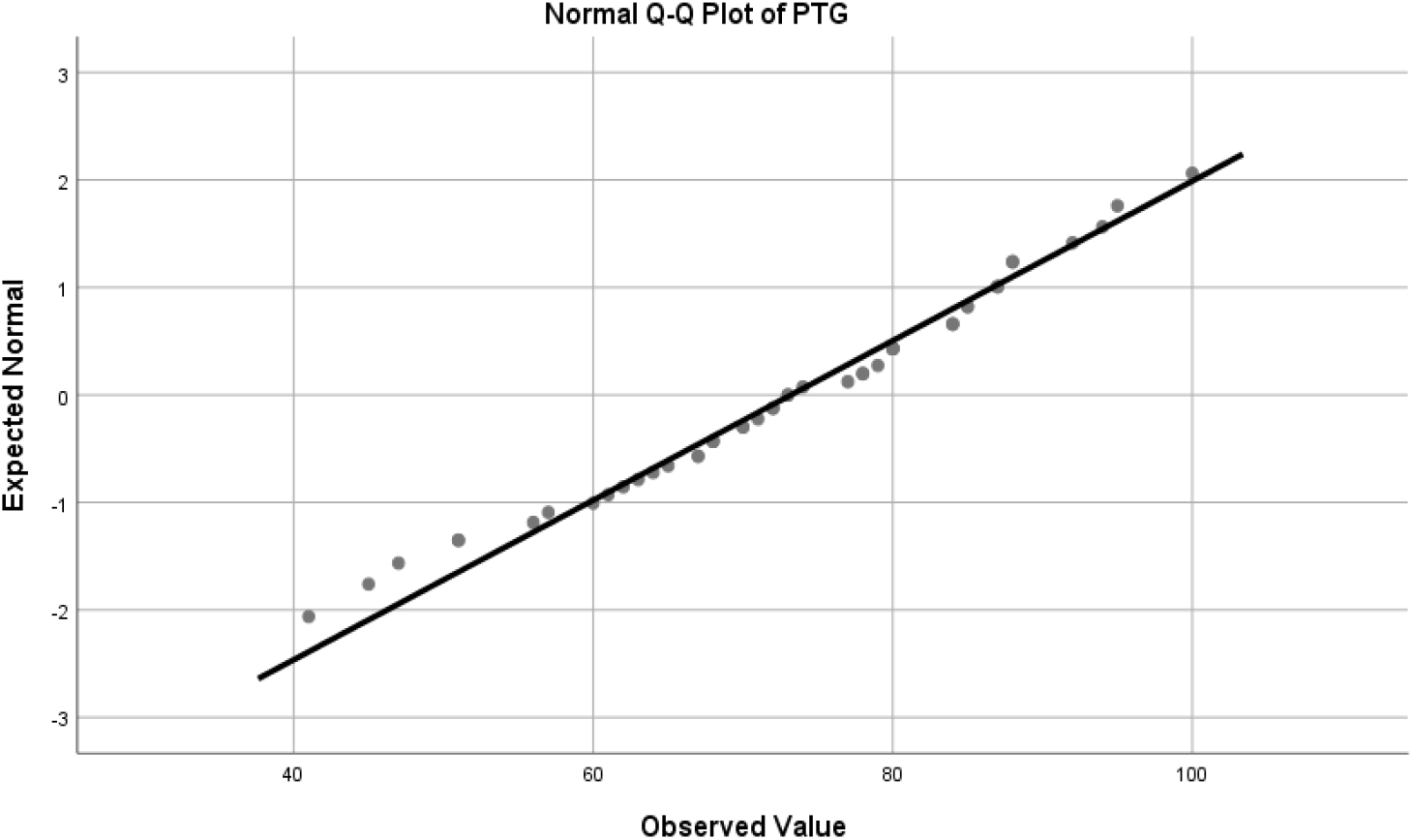
Quantitative diagram of normal quadratic growth variable. *Source: SPSS software output*

**Figure 2.**
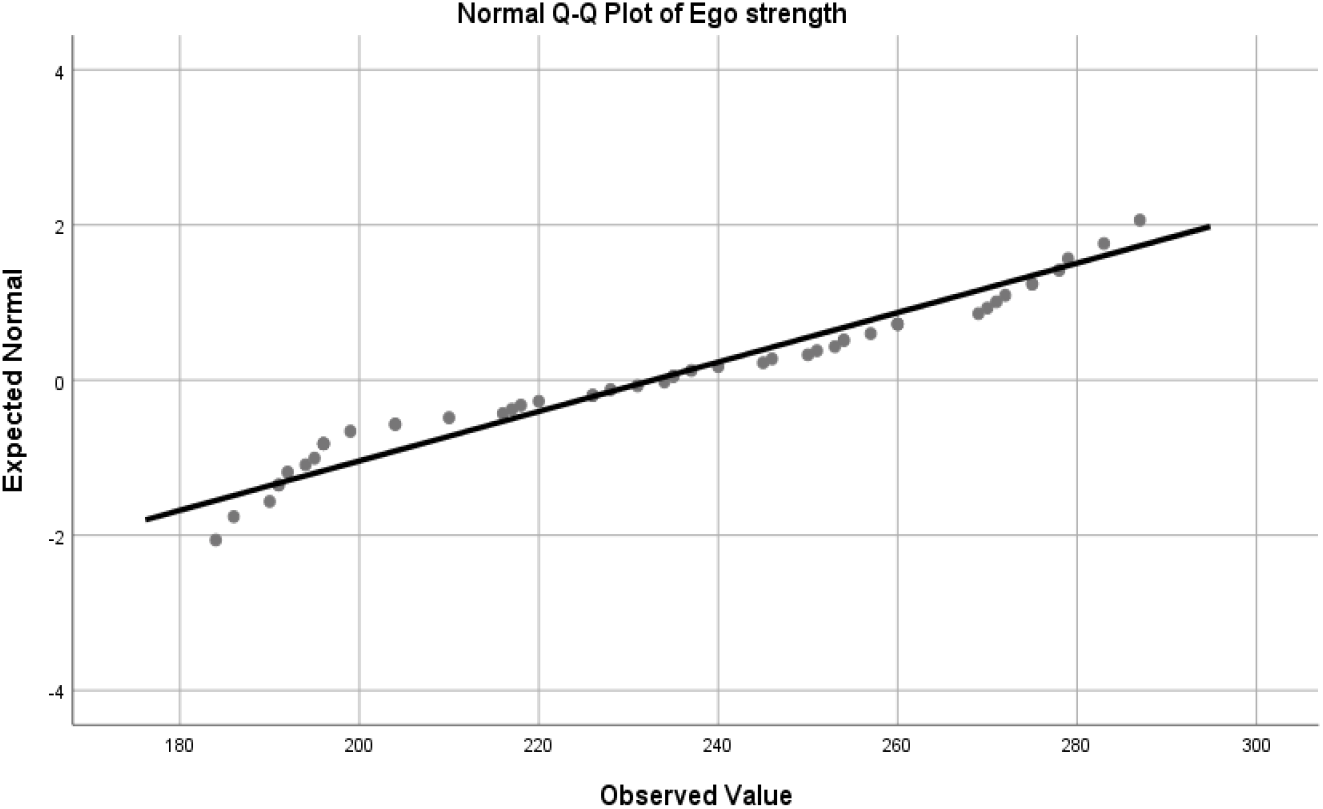
Graph of the normal multiplier of the ego strength variable. *Source: SPSS software output*

**Figure 3.**
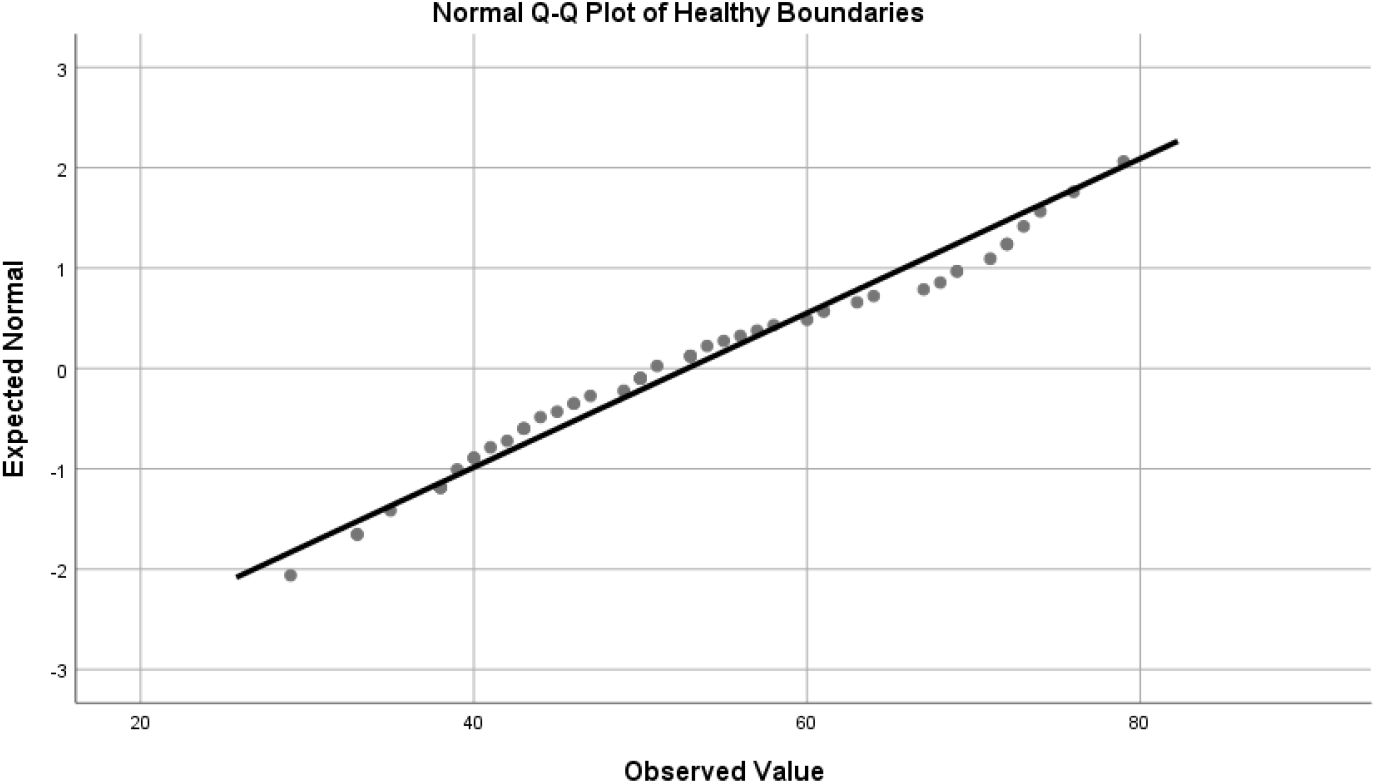
Normalmultiple diagram of a healthy bounedaries variable. *Source: SPSS software output*

**Figure 4.**
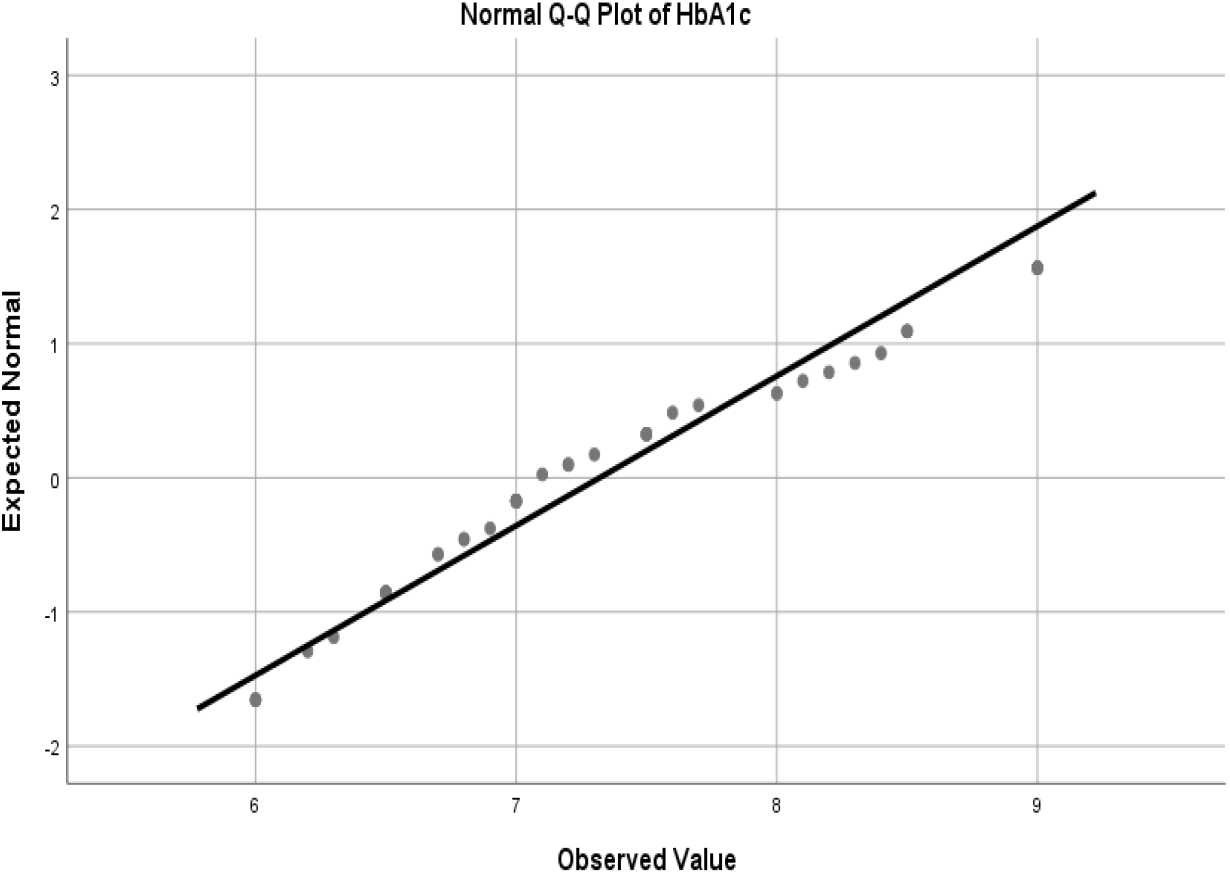
HbA1c Variable Normal Quantity Chart. *Source: SPSS software output*

**Figure 5.**
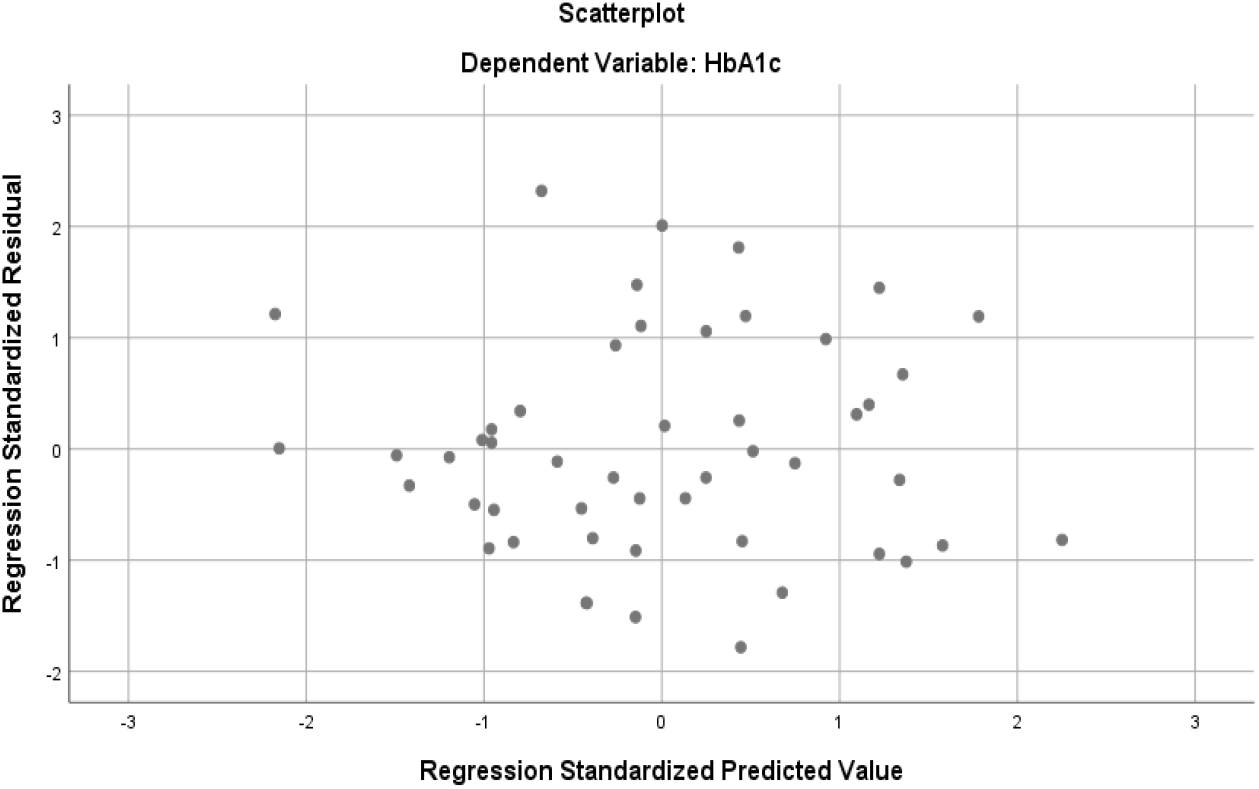
Residual distribution diagram and predicted values. *Source: SPSS software output*

**Figure 6.**
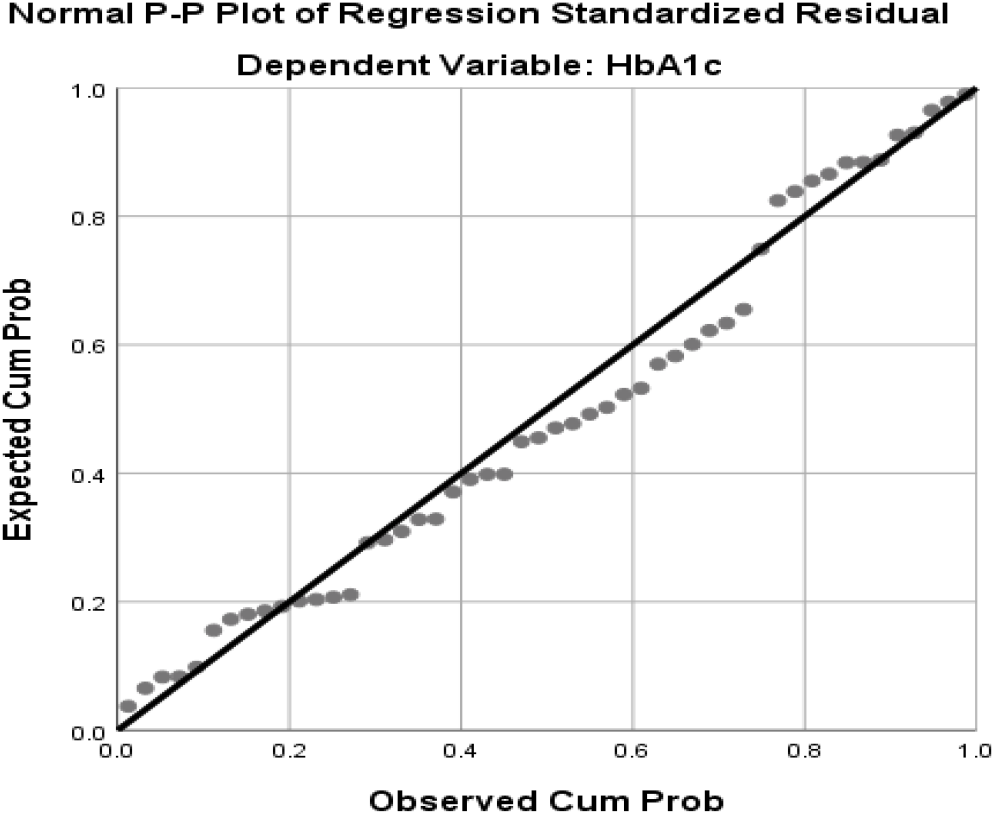
Normal multiple quadratic diagrams for residuals. *Source: SPSS software output*

### Watson Camera Test

After examining the regression assumptions, using multiple regressions, the predictive power of “HbA1c” was examined by each of the independent variables “post-traumatic growth”, “ego strength” and “healthy boundaries”. The regression model is as follows:

For the model whose dependent variable is “HbA1c”, according to Table 7, because the significance value is less than 0.05 (0.024) and the value of the F test statistic are 3.434, and then the fitted regression model is significant. In Table 6, the coefficient of determination between the independent variables “post-traumatic growth”, “ego strength” and “healthy boundaries” and the dependent variable “HbA1c” is 0.183, which means that the independent variables expresses about 18% of the variance of the dependent variable “HbA1c”.

**Table 6.** Summary of standard regression model

| Variables Entered/Removed<sup>a</sup> |  |  |  |
| --- | --- | --- | --- |
| Model | Variables Entered | Variables Removed | Method |
| 1 | HB<br>, PTG<br>, PIES | . | Enter |

a. Dependent Variable: HbA1c

b. All requested variables entered.
| Model Summary<sup>b</sup> |  |  |  |  |  |
| --- | --- | --- | --- | --- | --- |
| Model | R | R Square | Adjusted R Square | Std. Error of the Estimate | Durbin-Watson |
| 1 | .428<sup>a</sup> | .183 | .130 | .83612 | 1.136 |

a. Predictors: (Constant), PTG, PIES, HB
b. Dependent Variable: HbA1c

**Table 7.**
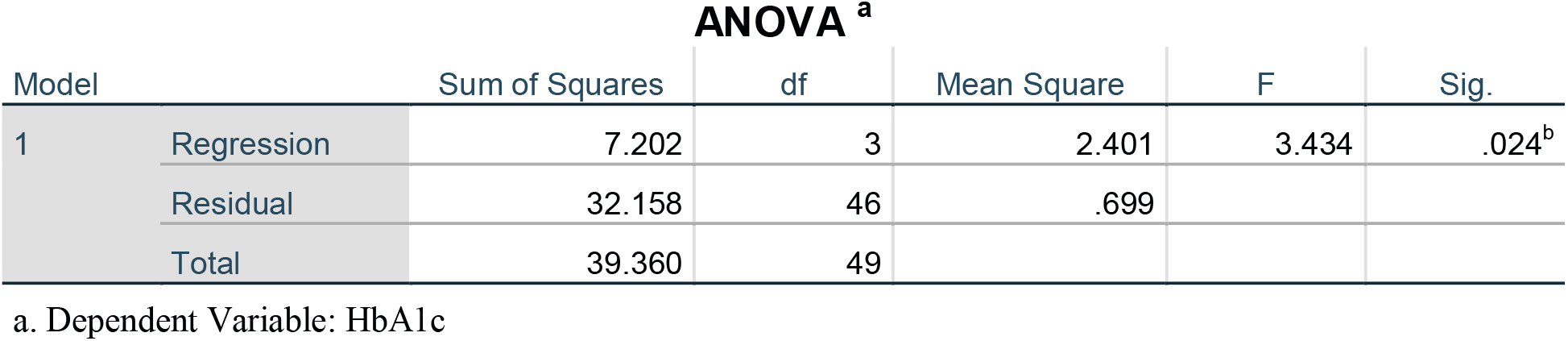
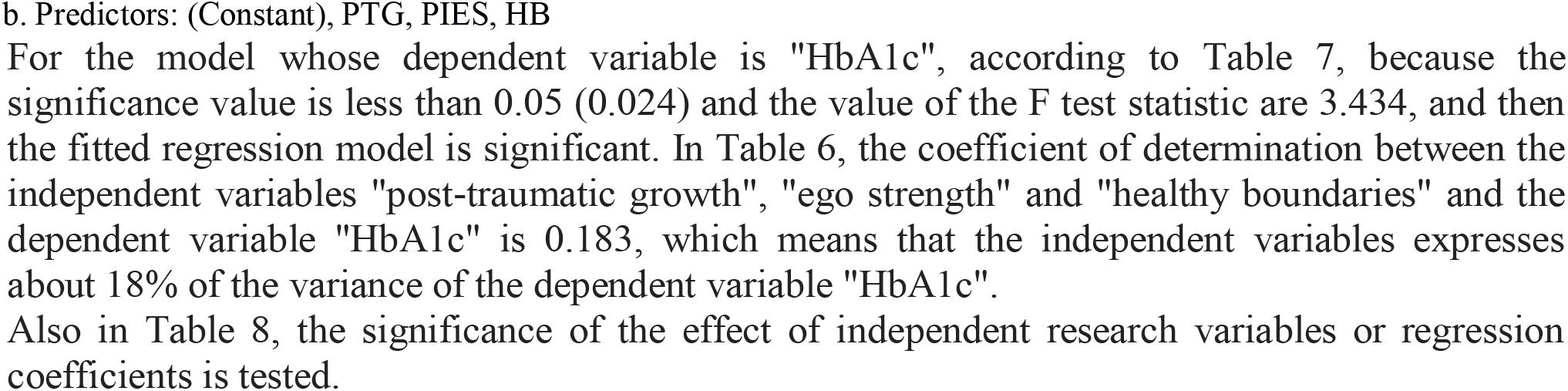
Significance test of standard regression model

| ANOVA<sup>a</sup> |  |  |  |  |  |  |
| --- | --- | --- | --- | --- | --- | --- |
| Model |  | Sum of Squares | df | Mean Square | F | Sig. |
| 1 | Regression | 7.202 | 3 | 2.401 | 3.434 | .024<sup>b</sup> |
|  | Residual | 32.158 | 46 | .699 |  |  |
|  | Total | 39.360 | 49 |  |  |  |

a. Dependent Variable: HbA1c
b. Predictors: (Constant), PTG, PIES, HB

Also in Table 8, the significance of the effect of independent research variables or regression coefficients is tested.

**Table 8.** Standard regression model coefficient

| Coefficients<sup>a</sup> |  |  |  |  |  |  |  |  |
| --- | --- | --- | --- | --- | --- | --- | --- | --- |
| Model |  | Unstandardized Coefficients |  | Standardized Coefficients |  |  | Collinearity Statistics |  |
|  |  | B | Std. Error | Beta | t | Sig. | Tolerance | VIF |
| 1 | (Constant) | 5.342 | 1.336 |  | 4.000 | .000 |  |  |
|  | PTG | -.023 | .009 | -.345 | -2.432 | .019 | .882 | 1.134 |
|  | PIES | .011 | .004 | .374 | 2.385 | .021 | .721 | 1.388 |
|  | HB | .022 | .010 | .322 | 2.163 | .036 | .803 | 1.245 |

a. Dependent Variable: HbA1c

For the independent variables “post-traumatic growth”, “ego strength” and “healthy boundaries”, given the significant value associated with them, which is less than 0.05, we can say that the independent variables “post-traumatic growth”, “strength” Ego and healthy boundaries have a significant effect on the prediction of the dependent variable HbA1c. Finally, according to the results of Table 8, the regression model of the research is as follows:

## Discussion

Because diabetes is very important, we need to identify people at risk for this disease. Since, ‘Ego boundaries’ is an accepted concept in psychotherapeutic circles. Building on work by Freud, Federn could be seen as the father of the concept as it is currently accepted by psychology and psychiatry. Initially, ego boundaries form the point at which the infant’s control of his world ceases (Federn, 1952b, p. 331).Since, A person without sufficient ego strength or healthy ego boundaries does not know what they want in life (E. Allers, personal communication, May 20, 2014), or if they do, they lack the assertiveness to attain their own goals, and are at the mercy of becoming subjugated to the more assertive wills of others, and according to the results of this study and the relationship between HbA1c blood sugar levels and ego strength, due to stress and insomnia can be concluded that ego strength is also effective in predicting and controlling HbA1c blood sugar levels.

A major cause of stress, overwhelm & burnt-outedness is the habit of ignoring the yearnings of our own True Selves while saying “yes” to other people’s requests (or demands) for our time, money, actions, even beliefs. In general, Healthy boundaries are those boundaries that are set to make sure mentally and emotionally you are stable.

As we know, Stress affects diabetes and blood sugar and Post-traumatic stress disorder (PTSD) is a psychiatric disorder that can occur in people who have experienced (directly or indirectly) or witnessed a traumatic event. It includes symptoms such as intrusion, avoidance, numbing, and hyper-arousal. Post-traumatic stress symptoms (PTSS) are often considered the most common negative psychological reactions in the aftermath of trauma.

In the other hand, Post-trauma growth (PTG) is defined as mastering a previously experienced trauma, perceiving benefits from it, and developing beyond the original level of psychological functioning (Tedeschi, Park, & Calhoun, 1998). According to the result obtained from the third hypothesis, there is a positive and significant relationship between predict blood sugar regulation and on posttraumatic growth (PTG) in diabetic patients.

The findings indicate a significant correlation between hyperglycemia and health boundaries, ego strength and post-traumatic growth. This means that controlling and recognizing the boundaries of mental health and post-traumatic emotions prevents high blood (HbA1c) sugar and Type 2 diabetes. It is recommended that future researchers:

-Review trauma in patients with diabetes what do they have, if it is the disease itself or other stressful events.
-Check the difference in HbA1c levels of the subjects before and after learning the boundaries, deeper research to determine whether the gene expression will change with teach the boundaries and stress control in the genome of an individual with a history of diabetes who may have inherited.
-Investigate the problems of diabetics economically, politically and socially.
-Also examine the neighborhood in which they live.
-Form groups to work on, people how to learn healthy boundaries and to dealing with stressful events and the formation of PTG houses in order to train post-traumatic growth to deal with high blood sugar and HbA1c.
-Organizing training classes for individuals, families and work environments.
-To deal with stressful events is suggested teaching life skills, strengthening the power of the ego and recognizing the boundaries of health to children.

## Conclusion

This study highlights the role and important of ego boundary, healthy boundaries and post trauma growth for researchers and psychologists. Since health boundaries, ego strength and post-traumatic growth play an essential role hyperglycemia. This leads us to realize that the effect of psychosocial health such as ego boundary, healthy boundaries and post trauma growth can be controlling and recognizing the boundaries of mental health and post-traumatic emotions prevents high blood (HbA1c) sugar and Type 2 diabetes.

## Data Availability

All data referred to in the manuscript are available and the resources are provided in the reference section.

